# The impact of keeping a religious beard in the COVID-19 pandemic: an online cross sectional survey study exploring experiences of male medical healthcare professionals

**DOI:** 10.1101/2020.11.01.20224022

**Authors:** Amer Hamed, Asam Latif, Mohammad Haris, Sufyan Patel, Muhammad I Patel, Syed Abdur Rahman Mustafa, Omeair Khan, Ahmad Shoaib, Salman Waqar

## Abstract

There has been a disproportionate effect on individuals from black Asian & Minority Ehnic (BAME) in the UK in Coronavirus disease (COVID-19) pandemic, especially in the NHS staff. Many of them have been asked to remove their beard to be eligible to do the fit test which can have negative implications on their spiritual, psychological & emotional wellbeing. This paper surveyed the responses of 469 healthcare professionals (HCPs) with beards regarding the challenges they face in regard to personal protective equipment (PPE), mask fit testing and attitude of employers & colleagues.

Professional discrimination through fit testing rejection, unavailability or inadequate PPE and the pressure to shave their beard are unpleasant outcomes of this pandemic for some of the NHS staff. NHS trusts & hospitals need to adjust their practices to include staff with beard in their COVID-19 arrangements.

## Introduction

The COVID-19 pandemic has required unprecedented adaptation of work systems and practices globally. (1) Research indicates that there has been a disproportionate effect on individuals from BAME backgrounds, particularly affecting key healthcare workers. Indeed, the first 11 doctors who lost their lives to this pandemic belonged to BAME communities. (2)

There has been much debate on the reasons why minority groups have been adversely affected. One reason is the availability and use of PPE for HCPs. There are multiple guidelines regarding appropriate PPE to protect clinicians and patients from the spread of COVID-19. For HCPs, when dealing with aerosol-generating procedures (AGPs) (e.g. cardiopulmonary resuscitation), the guidelines are unanimous in instructing that a higher level of PPE is required. This is generally a filtering facepiece (FFP3) mask or a powered air-purifying respirator (PAPR) hood. (3-5) The FFP3 mask requires fit testing to ensure a secure seal is formed to provide adequate protection. (6) Fit testing for respiratory protective equipment (RPE) has been a requirement in the USA since 1969 and in the UK since 1999. (7-8) The purpose of fit testing is to verify that the specific RPE used has the potential to provide adequate protection for the individual. This means it cannot guarantee adequate protection at all times as this depends on the user ensuring they always fit the mask correctly. (6)

Unfortunately, fit testing poses several challenges. (9-10) Logistically, it is difficult to fit test all clinical staff within a short period of time and urgency of testing in a pandemic makes it problematic. (11) There are also concerns about wastage of several FFP3 masks and indispensable PPE kit during this testing in the middle of a pandemic. (12) Nevertheless, many individuals are unable to pass the fit testing due to their facial anatomy limiting a perfect seal, and others fail due to the presence of facial hair (or beard). (13) The PPE protection is, however, essential in fighting this pandemic and hence making it imperative to consider the respirator hood. The hood is a worthwhile, but costly alternative that provides effective protection and is not affected by the aforementioned issues. (3)

Many clinicians have been asked to remove their beard during the COVID-19 pandemic. A beard may be kept as a fashion trend for some people, and therefore shaving it is then an understandable and duty-bound sacrifice. However, for many individuals, a beard is a sacred aspect of religious practice. (14) This includes many religions such as Islam, Judaism, Sikhism and some sects within traditional Christianity. Many followers of these religions consider their beards to be an integral part of their personal and religious identity. (14) There is anecdotal evidence to suggest that male healthcare professionals have been asked to shave their beards in order to be eligible. (15) Requesting the removal of their beards in order to be eligible for fit testing could have huge negative implications on their psychological, spiritual, and emotional wellbeing as this is considered to be a religious requirement for many faith groups. (16)

### Primary research objectives

- To ascertain what challenges healthcare professionals who wore religious beards faced in regards to PPE, mask fit-testing and attitudes of employers and colleagues.
- To estimate the proportion of men who kept religious beards that subsequently trimmed or shaved due to circumstances during the COVID-19 pandemic.
- To estimate the proportion of men who wore a religious beard that faced challenges in their workplace.
- To gather the views and opinions on why men who kept religious beards trimmed or shaved, or, made no changes.

## Methods

### Study design

This cross-sectional study was conducted using online social media tools from April to May 2020. Social networking sites (Facebook, Linkedin, Twitter and Whatsapp) were used to distribute the survey and adhere to COVID-19 guidelines. Participants were assured that the information they provide would be kept anonymous and confidential. The survey was created using Google Forms and a link to participate was distributed.

A qualifying question was asked at the beginning of the survey and completion of the questionnaire implied informed consent. Items for the survey were derived from the literature and through thorough discussions with experts and colleagues within the profession. The instrument contained a total of 36 questions. A variety of question types were used including Likert-type scales with responses on a ten-point scale ranging from “strongly agree” to “strongly disagree”, closed-ended questions and open-ended questions. The questionnaire consisted of three sections with the final section requiring participants to provide their socio-demographic details. This study was intended to be a descriptive study thus the questionnaire used was not validated.

### Patient and public involvement statement

No patients were involved in this study. Any self-identifying healthcare professional who wore a religious beard was eligible to participate. For the purposes of this study we defined a healthcare professional as any individual working in either a primary or secondary healthcare setting in a public facing role.

### Pilot Testing

The questionnaire was piloted by both academics and lay personnel. The aim of the pilot testing was to ascertain time taken to fill out form, in what order questions were asked, whether questions were understood as intended and if the questionnaire was user friendly. The feedback alongside responses from the pilot survey were then discussed by the study team and relevant changes were made to the questionnaire. The pilot responses were then removed before the finalised questionnaire was uploaded to Google Forms.

### Analysis Plan

The data was downloaded from Google Forms and inputted into a Microsoft Excel spreadsheet. After cleaning and consistency checks, the data was transferred to the Statistical Package for Social Sciences (SPSS) version 25 and was again checked for consistency and correctness. Descriptive analysis using frequency distributions, confidence intervals, distribution (mean etc), was used to analyse the data.

### Ethical consideration

The self-complete questionnaire was optional and therefore had few ethical implications. Consent was implied upon completion of the questionnaire. Following enquiry with the Health Research Authority (HRA), approval was not deemed necessary. Rather, approval from the BIMA ethics committee was sufficient. Adherence to ethical standards are maintained by informed consent, anonymising the data & provision of study information.

## Results

### Demographic profile of participants

A total of 469 healthcare professionals participated in this survey. The number of respondents varied for individual questions in this survey and they are clearly detailed in Tables 1 – 4.

**Table 1:** Demographic information

| **Survey Question** | **No of Participants** | **Result** |
| --- | --- | --- |
| **Q 1: Age**<br>(Mean) (SD) | 462 | 39 (10)<br>Range: 20 -69 |
| **Q 2: How long have you been working in healthcare?** |  |  |
| Under 5 year: | 465 | 120 (26%) |
| 5-10 years | 465 | 80 (17%) |
| Over 10 years | 465 | 265 (57%) |
| **Q 3: Ethnicity** |  |  |
| Asian/Asian British | 456 | 383 (84%) |
| Middle Eastern | 456 | 39 (9%) |
| White | 456 | 10 (2%) |
| Black | 456 | 11 (2%) |
| Any other ethnic group | 456 | 11 (2%) |
| Mixed | 456 | 2 (0.44%) |
| **Q 4: Country of Birth?** |  |  |
| UK | 447 | 169 (38%) |
| India/ Pakistan/Bangladesh | 447 | 184 (41%) |
| Africa | 447 | 44 (10%) |
| Middle east | 447 | 18 (4%) |
| Other | 447 | 32 (7%) |
| **Q 5: Country of work** |  |  |
| UK | 436 | 427 (94%) |
| Outside UK | 436 | 27 (6%) |
| **Q 6: City of Work** |  |  |
| London | 436 | 67 (15%) |
| Manchester | 436 | 33 (8%) |
| Leicester | 436 | 30 (7%) |
| Birmingham | 436 | 22 (5%) |
| Blackpool | 436 | 16 (4%) |
| All Others | 436 | 268 (61%) |
| **Q 7: Your role in healthcare?** |  |  |
| Doctor | 464 | 372 (80%) |
| Allied healthcare worker | 464 | 34 (7%) |
| Nurse | 464 | 9 (2%) |
| Others | 464 | 49 (11%) |
| **Q 8: Place of work** |  |  |
| Hospital | 469 | 390 (83%) |
| General practice |  | 36 (8%) |
| Other places |  | 43 (9%) |
| **Q 9: What contract do you have with your board?** |  |  |
| Permanent/substantive | 467 | 258 (55%) |
| Training | 467 | 107 (23%) |
| Locum/Bank | 467 | 66 (14%) |
| Part time | 467 | 16 (4%) |
| Other | 467 | 20 (4%) |

| **Q 10: Religion** |  |  |
| --- | --- | --- |
| Muslim | 467 | 437 (94%) |
| Sikh | 467 | 18 (4%) |
| Jewish | 467 | 6 (1%) |
| Other/Not to say | 467 | 5 (1%) |
| **Q 11: Did you get suspected or confirmed Covid-19?** |  |  |
| Yes | 467 | 119 (25%) |
| No | 467 | 348 (75%) |

**Table 2:** General Questions

| **Survey Question** | **No of Participants** | **Result (% , 95% CI)** |
| --- | --- | --- |
| **Q 1: Do you think your beard represents your religious identity?** |  |  |
| Yes | 469 | 444 (95%)<br>(92-96) |
| No | 469 | 25 (5%)<br>(4-8) |
| **Q 2: Do you know that “religion or belief” is one of nine protected characteristics under the Equality Act 2010?** |  |  |
| Yes | 469 | 332 (71%)<br>(66 – 75) |
| No | 469 | 137 (29%)<br>(25-34) |
| **Q 3: To what extent do you feel pressurised by your colleagues, department or Trust to trim/shave your beard during the current pandemic (rate it from 1-10)?** |  |  |
| 1-4 | 469 | 214 (46%)<br>(41-50) |
| 5-7 | 469 | 116 (25%)<br>(21 -29) |
| 8-10 | 469 | 139 (29%)<br>(26 – 34) |
| **Q 4: Did you get a mask fit test?** |  |  |
| Not offered a test due to beard | 469 | 168 (36%)<br>(32 – 40) |
| Did not being bother fit test due to beard | 469 | 85 (18%)<br>(15 – 22) |
| Failed due to beard | 469 | 52 (11%)<br>(8 – 14) |
| Passed only after shaving | 469 | 53 (11%)<br>(9 – 15) |
| Passed with beard | 469 | 27 (6%)<br>(4 – 8) |
| Passed only after trimming | 469 | 21 (5%)<br>(3 – 7) |
| Not offered/Other reasons | 469 | 63 (13%)<br>(10 – 17) |
| **Q 5: Did your Trust provide you any alternative PPE (e.g. Hood) due to your beard?** |  |  |
| Yes | 469 | 103 (22%)<br>(18 – 26) |
| No | 469 | 296 (61%)<br>(56 – 65) |
| Not applicable | 469 | 80 (17%)<br>(14 – 21) |
| **Q 6: If you failed fit test or were not provided with alternative PPE, were you offered alternate form of work?** |  |  |
| Yes, advised not to perform high risk procedures | 469 | 44 (9%)<br>(7 – 12) |
| Yes, offered to work in non-clinical area | 469 | 14 (3%)<br>(2 – 5) |
| Yes, offered to work from home | 469 | 14 (3%)<br>(2 – 5) |
| Yes, any other alternate form of work | 469 | 11 (2%)<br>(1 – 4) |
| No alternative option of work was given | 469 | 213 (45%)<br>(41 – 50) |
| Did not fail fit test or was offered alternative PPE | 469 | 173 (37%)<br>(33 – 41) |
| **Q7: If you failed fit test or were not provided with alternative PPE, did you consider buying your own PPE?** |  |  |
| Yes, I considered and used my own PPE | 469 | 63 (13%)<br>(11 – 17) |
| Yes, I considered buying my own PPE but was not allowed to use own PPE | 469 | 25 (5%)<br>(4 – 8) |
| Yes, have considered buying my own PPE but could not find or afford it | 469 | 95 (20%)<br>(17 – 24) |
| No, I didn't consider it | 469 | 160 (34%)<br>(30 – 39) |
| Did not fail fit test or was offered alternative PPE | 469 | 126 (27%)<br>(23 – 31) |
| **Q 8: Does your Trust have a policy on PPE and beards?** |  |  |
| Don't know | 469 | 246 (52%)<br>(48 – 57) |
| No | 469 | 143 (30%)<br>(26 – 35) |
| Yes | 469 | 80 (17%)<br>(14 – 20) |
| **Q 9: Were you given the opportunity to speak to a senior clinician or manager in your Trust to discuss PPE and beard issues?** |  |  |
| Yes | 469 | 124 (26%)<br>(23 – 30) |
| No | 469 | 345 (74%)<br>(69 – 78) |
| **Q 10: Were you given any written instructions to trim or shave your beard?** |  |  |
| Given written instruction | 469 | 46 (10%)<br>(7 – 13) |
| Given verbal instruction and was denied written instruction | 469 | 19 (4%)<br>(3 – 6) |
| Given verbal instruction and I did not ask for written instruction | 469 | 150 (32%)<br>(28 – 36) |
| Was not asked to trim or shave my beard | 469 | 254 (54%)<br>(50 – 59) |
| **Q 11: Did you shave or trim your beard due to Covid-19?** |  |  |
| Yes | 469 | 176 (38%)<br>(33 – 42) |
| No | 469 | 293 (62%)<br>(58 – 67) |

**Table 3:** Participant who have not changed beard

| Survey Question | No of Participants | Result (%, 95% CI) |
| --- | --- | --- |
| **Q 1: How confident you feel that you can retain beard during this pandemic (rate form 1-10)?** 1 – Not sure at all, 10 – fully confident to retain my beard |  |  |
| 1-4 | 293 | 31 (11%)<br>(8 – 15) |
| 5-7 | 293 | 56 (19%)<br>(15 – 24) |
| 8-10 | 293 | 206 (70%)<br>(65 – 75) |
| **Q 2: Do you feel that you have adequate PPE to protect yourself with a beard?** |  |  |
| Yes | 293 | 130 (44%)<br>(39 – 50) |
| No | 293 | 163 (56%)<br>(50 – 61) |
| **Q 3: Do you feel that your colleagues are supportive for you to keep beard (rate from 1-10)? 1 - Not all supportive, 10 - fully supportive** |  |  |
| 1-4 | 293 | 63 (22%)<br>(17 – 27) |
| 5-7 | 293 | 117 (40%)<br>(34 – 46) |
| 8-10 | 293 | 113 (38%)<br>(33 – 44) |
| **Q 4: 4. Do you feel a need for change in working pattern or place due to your beard?** |  |  |
| Yes | 293 | 96 (33%)<br>(28 – 38) |
| No | 293 | 197 (67%)<br>(62 – 72) |
| **Q 5: Did your department make any changes in work pattern to accommodate your beard?** |  |  |
| Yes | 293 | 39 (13%)<br>(10 – 18) |
| No | 293 | 254 (87%)<br>(82 – 90) |
| **Q 6: 6. Do you feel threatened to lose your job for keeping a beard during this pandemic?** |  |  |
| Yes | 293 | 48 (16%)<br>(12 – 21) |
| No | 293 | 242 (83%)<br>(78 – 87) |
| Yes, already lost my job due to this issue | 293 | 3 (1 %)<br>(0.3 – 3) |

**Table 4:** Participant who trimmed or shaved beard

| Survey Question | No of Participants | Result (%, 95% CI) |
| --- | --- | --- |
| **Q 1: 1. The reason for trimming/shaving? (Participants can select more than one option)** |  |  |
| Voluntary | 176 | 67 (38%)<br>(34 – 42) |
| Perceived risk of getting illness | 176 | 88 (50%)<br>(46 – 54) |
| Pressure from employer | 176 | 48 (27%)<br>(23 – 31) |
| Lack of alternative PPE | 176 | 102 (58%)<br>(53 – 63) |
| Threat to lose job | 176 | 15 (8%)<br>(4 – 13) |
| Other | 176 | 27 (15%)<br>(11 – 19) |
| **Q 2: Was the opinion and guidance of a clinician, manager or faith leader imposed on you?** |  |  |
| Yes | 176 | 37 (21%)<br>(16 – 28) |
| No | 176 | 139 (79%)<br>(72 – 84) |
| **Q 3: Do you feel guilt for trimming/shaving your beard?** |  |  |
| Yes | 176 | 107 (61%)<br>(53 – 68) |
| No | 176 | 69 (39%)<br>(32 – 47) |
| **Q 4: 5. Has trimming/shaving your beard affected your mental or emotional wellbeing?** |  |  |
| Yes | 176 | 74 (42%)<br>(34 – 49) |
| No | 176 | 102 (58%)<br>(50 – 65) |
| **Q 5: 6. Do you feel your religious identity has been violated?** |  |  |
| Yes | 176 | 88 (50%)<br>(43 – 57) |
| No | 176 | 88 (50%)<br>(43 – 57) |
| **Q 6: Do you feel that your civic or human rights have been violated?** |  |  |
| Yes | 176 | 73 (41%)<br>(34 – 49) |
| No | 176 | 103 (59%)<br>(51 – 66) |

Almost all of our cohort came from BAME backgrounds with the majority of respondents of Muslim faith. More than three quarters were doctors. The hospital workplace was the predominant place of work. More than half of the participants had permanent contracts. The majority of respondents were from Britain and London was the most common workplace (67, 15%).

The median age of respondents was 39 years (range 20 - 69). There were varying ethnicities of respondents with the vast majority coming from Black, Asian and Minority Ethnic (BAME) groups (446, 98%). Asian / British Asian ethnicity made up the bulk of the BAME group (383, 84%). Only a minority were of a white background (10, 2%) possibly accounted for by the significantly smaller proportion of white people sharing Muslim, Sikh or Jewish beliefs. The majority of respondents shared the Muslim faith (437, 94%) followed by the Sikh faith (18, 4%) then the Jewish faith (6, 1%).

The highest percentage of respondents had worked in the NHS for greater than 10 years (265, 57%). Doctors accounted for the majority of respondents (372, 80%) whilst nurses (9, 2%), allied healthcare workers (34, 7%) and other support staff (49, 11%) also responded. Places of work included hospitals (390, 83%), GP practices (36, 8%) and social care among others. The types of contracts varied with the majority having permanent posts (258, 55%) whilst trainees and locums accounted for 23% (107) and 14% (66) respectively.

Of our cohort 25% (119) of respondents by the 14th of May 2020 had confirmed positive for SARS-CoV-2 (COVID-19).

### The beard

From our cohort 444 (95%) respondents felt that the beard was part and parcel of their religious identity. Despite the majority of participants (332, 71%) knowing that ‘religion or belief’ was a protected characteristic under the Equality Act 2010, more than half (255, 54%) of respondents who have a beard for religious reasons felt moderately or highly pressured by their colleagues, departments or trusts to trim or shave their beards during the pandemic.

### FIT testing

The majority of respondents were not offered a FIT test for the FFP3 face mask due to wearing a beard (168, 36%) or they did not pursue getting one (85, 18%). 11% (52) of respondents failed the FIT test due to having a beard. Of those who passed the FIT test, 6% (27)passed with their beard, 5% (21) after trimming and 11% (53) after shaving. Almost three quarters of our respondents (345, 74%) were not given the opportunity to discuss their concerns regarding PPE and their religious observance of a beard with a senior clinician or a manager.

### Alternative PPE

Among the alternatives to the FFP3 mask are respirator hoods which do not require fit testing for use and can accommodate clinicians that fail FIT testing or wear beards. From our respondents that wore beards for religious observance, 61% (296) were not provided any PPE or were refused any alternatives. 22% were supported by their Trusts and alternate PPE was provided. Of those who were not provided with alternative PPE, 13% (63) purchased and used their own PPE, 20% (95) considered buying their own PPE but either couldn’t afford it or found an appropriate model and 5% (25) who considered buying their own PPE were not allowed to use it.

### Amended duties

Of those who failed FIT testing (either due to not being tested or actually failing the test) and were not offered alternative PPE; 45% (213) were not offered any alternative duties at work. In the same group, Only 9% (44) were advised to abstain from high risk procedures, and less than 5% (28) offered work in non-clinical areas or to work from home.

### Participants who changed their beards

From our respondents 38% (176) either trimmed or shaved their beard due to COVID-19. The majority who shaved or trimmed their beards cited lack of PPE (102, 58%) and perceived risk of becoming unwell (88, 50%) as the two main reasons. More than a quarter (48, 27%) felt pressured from their employer to remove their facial hair.

Shaving the beard affected the mental or emotional wellbeing of 42% (74) of respondents whilst 61% (107) felt guilty for doing so. Half of respondents felt their religious identity had been violated (50%, 88).

### Participant who retained their beards

From our respondents 62% (293) didn’t trim or shave their beard due to COVID-19. Of this group, More than half (163, 56%) feel they do not have adequate PPE available to protect themselves with a beard. The majority (262, 78%) felt moderately or highly supported by their colleagues to keep a beard for religious observance. However, 22% (63) experienced little support from their colleagues. A minority of departments (19, 13%) made changes to the work pattern of their employees who retained beards for religious observance. A handful of respondents (3, 1%) were sacked for refusing to shave their beards for religious observance.

## Discussion

The recent Public Health England (PHE) report details the impacts of COVID on BAME individuals and explores the effect of health inequalities within these groups. It emphasises that faith “provides an important foundation for communities’ resilience” highlighting the major role faith plays in helping an individual overcome such challenges. (17) Unfortunately, there have been reports of Islamophobia within the health service which undermines efforts to support people practicing their religion. (18) This study aimed to explore and propose reasonable solutions to the challenges faced by some HCPs who observe religious practice by keeping a beard. Investigating appropriate PPE for AGPs in clinical practice, our study is committed to determine a solution that protects religious identity whilst ensuring that the priority of saving lives is not compromised.

Many HCPs who keep a beard for religious reasons are from religious minority groups and so the possible discrimination may lead to a decrease in workforce diversity. The fit test processes currently do not appear to be consistent with the Human Rights Act. (19) Others have suggested that the lack of appropriate workwear has reduced opportunities for female Muslim women. (20) A costly but viable alternative to a fitted respirator mask is a respirator hood; this is a reasonable substitute that should be considered where religious identity issues are present. However, many HCPs are unaware of such alternatives and organisations may not be prepared to consider such costly options. (16, 21)

Workforce diversity helps to improve patient care. People from ethnic minority faces racial and professional discrimination during stages of their medical careers. (22) Muslims are the highest religious group to face such discrimination. (23) This discrimination was felt by the majority of healthcare professionals in a recent survey. (18)

Recent NHS new national guidance for NHS organisations in England published by the NHS employers suggested that the staff shave off their beards and facial hair to ensure protective face masks fit properly. (24)

Maintaining a religious beard comes with its challenges for the staff of the NHS in the current pandemic. Professional discrimination through fit testing rejection, unavailability or inadequate PPE and the pressure to shave your beard are unpleasant outcomes of this pandemic for NHS staff from primarily Muslim faith as the study shows.

Discrimination against staff is linked to poorer outcomes for patients, and little progress has been made in the last two decades on this front. Our findings are consistent with previous research which demonstrated discrimination faced by Muslim women with respects to dress codes in theatre and in hospital ward environments.

Furthermore, research into PPE provision in BAME frontline staff during the Covid-19 pandemic also showed that Muslims reported a higher rate of being reprimanded for reporting PPE issues in the workplace, reported higher rates of Covid-19, and had the most difficulty accessing PPE. (25)

NHS staff from the faith groups need to feel accommodated, supported & empowered by their colleagues and trusts. Maintaining diversity in the workplace is important to improve the NHS care as well as its benefits to the society as a whole. NHS trusts & hospitals need to adjust their practices to include staff with beard in their Covid-19 arrangements.

Staff from faith groups who are subjected to such discrimination need to approach the diversity personnel in their trusts or seek help & support from their faith group.

### Implications

The findings from this study shows that not enough consideration was given to the importance of the beard to health care staff, by not accommodating those who have it through failure to supply alternative PPE or direct the staff to an alternative type of work. In spite of this, staff still have faith in the health system to accommodate their religious needs.

### Strengths and Limitations

For the purpose of this cross-sectional study, social media platforms were used for disseminating the survey. The anonymity of respondents in this survey would facilitate honest reflections and the ability to speak freely on their sensitivities with regards to the survey. From the dataset gathered, it is clear that the majority of respondents were Muslims and there was relatively little representation of Jewish and Sikh respondents. It is also difficult to objectively assess how representative this survey would be of the NHS given that respondents were responding to social media advertisement to participate in the survey and some respondents were working in other countries. Furthermore, the sample was self-selected thus it may not equally reflect opinions and practices across the NHS.

The cross-sectional nature of the study doesn’t take into account changes in opinions of healthcare staff and the practices across different hospitals over time to adapt to the attitudes of staff, the needs of clinicians and the effect on practice as the pandemic eases. On the other hand, this descriptive study does look at how practices within the NHS were around the peak of the pandemic and highlights important issues which may have not been considered yet, which significantly affected healthcare workers in a practical and spiritual sense.

### Further research

Further research will be needed to expand more on these important issues and challenges facing healthcare professionals who have a beard in regards to PPE, mask fit-testing and attitudes of employers and colleagues.

## Data Availability

All data available on the study google form.

## Acknowledgements

We would like to acknowledge all the participants who generously gave their time to complete the questionnaire. We extend a special thanks to Dr Mohammed Javed Iqbal, Dr Mohammed Bilal Patel who contributed significantly to the success of the study, Sharjeel Abu Ameerah and Yahya Hassan for their contribution in the study, Fayyaz Khan for reviewing the introduction. We also give thanks to the British Islamic Medical Association (BIMA) for their support especially Dr Arshad Latif who helped in forming the group of people interested in this subject.

## Supplements

